## Additional File 1 for "Predicting the causative pathogen among children with pneumonia using a causal Bayesian network"

#### Additional File 1. Survey questions
#### Survey 1 - distributed prior to Workshop 1.

Please rate each variable within each submodel (A, B, etc), on a scale of 1 - 10 regarding their relevance in differentiating 1) causative pathogens, and 2) clinical severity/ risk of complications, 1 as minimal level of relevance and 10 as the highest.

| Submodel; proposed high level structure | Key questions of interest for each submodel | Variables | Rate importance within each submodel (A, B, etc.) | | |
|---|---|---|---|---|---|
| | | | Causative pathogens (e.g., bacterial vs non-bacterial) | Clinical severity/risk of complications | Comment |
| *A. Background factors* | **1.** How background factors influence exposure to different organisms; i.e., where the bugs are in the host's environment. **2.** How they influence one's susceptibility to disease upon pathogen exposure; i.e., the host defense aspect. **3.** How different organisms reside/ colonise on different parts of body; bug-host interaction. **4.** How they influence the decision to conduct certain clinical/ lab investigations, and subsequent management of patient. | Age<br>Gender<br>Ethnicity/ Indigenous status<br>Country of birth<br>Gestation, weeks<br>Language used at home<br>*Comorbidity*<br>*Immunisation status*<br>*Exposure to pathogens*<br>Past antibiotic use<br>*Social economic factors*<br>*Site of colonisation*<br>*Colonising organism*<br>Density of colonisation<br>*Management prior to visit*<br>Please specify other variable/s if not listed above | | | |
| ***B. Current infection episode*** | **1.** How bugs interact with each other and the host's immune system to develop infection episode; i.e., understand the natural history of disease/ the fight process between the bug and host immune system. **2.** How such processes differ by sites (e.g., certain strains are better at invading into the lung?) **3.** Co-infections? Multi-sites? **4.** How bug-host interaction manifests as symptoms and signs. | *Infecting pathogen/s*<br>*Site/s of infection*<br>*Immune and inflammatory responses*<br>Empyema<br>Please specify other variable/s if not listed above | | | |
| ***C. Symptoms and signs*** | **1.** How symptoms and signs can drive clinical and lab investigations. **2.** How they interact and get reported. **3.** How infection-induced changes (e.g., elevated bio markers) can be detected as clinical and lab measures. **4.** How symptoms and signs drive decision for management. **5.** Should (and which) symptoms be evaluated in the same way at the initial point-of-care vs post-management (i.e., outcome evaluation) | *Systemic (non-respiratory) symptoms*<br>*Respiratory abnormalities*<br>*Cadiovascular related*<br>*Symptoms specific for small babies*<br>Please specify other variable/s if not listed above | | | |
| ***D. Investigations*** | **1.** Understand the clinical and lab workflow for testing including factors driving decisions on which investigations to perform; e.g., resource, timing, cost, etc. **2.** What is the aim of a given investigation; e.g., detection of organism (and its quantity), detection of immune response. **3.** How testing results should be interpreted to inform diagnosis such as causative pathogen and disease severity; e.g., threshold for abnormality (potentially age-specific), false negatives, false positives, multiple growths, contamination rates. | *Haematology*<br>*Inflammatory biomarkers*<br>*Type/ site of specimen*<br>*Method of pathogen detection*<br>*Imaging*<br>Detected quantity (density) of organism/s<br>Contamination<br>*AMR-related*<br>Urinalysis<br>Please specify other variable/s if not listed above | | | |
| ***E. Management*** | **How does the knowledge of causative pathogen or severity influence management decisions. 1.** How diagnosed (or, in the future, model-predicted) disease (causative pathogen/s, sites, severity) drives the choice of antibiotics (route, spectrum, dose). **2.** How evidence and diagnoses influence management (e.g. supplementary oxygen, admission, discharge). | Use of antibiotic<br>Spectrum of antibiotic<br>Route of antibiotic<br>Dose of antibiotic<br>*Other medications*<br>Supplementary oxygen<br>Intra-venous fluid<br>Physio/ airway clearance therapy<br>Surgery<br>*Disposition*<br>Please specify other variable/s if not listed above | | | |
| ***F. Outcomes*** | **How does the knowledge of causative pathogen or severity influence patient outcomes. 1.** Which endpoints to use for evaluating patient outcome. **2.** Where these endpoints are located in the causal pathway of the disease; e.g., does symptom clearance equal eradication of pathogen? **3.** How they may be evaluated differently for different patient subgroups. | Re-presentation (to GP, hospital)<br>Modification of antibiotics<br>Quality of life<br>*Function related*<br>*Symptom related*<br>*Psychological*<br>*Complications*<br>Please specify other variable/s if not listed above | | | |

#### Survey 2 – distributed prior to Workshop 2

**Variation in detection pathways & organisms**

Here is a list of organisms that were sought and detected in the pneumoWA project. There were two pathways for obtaining a detection result: study and clinic. Quantification was performed for some organisms via the study pathway. More detailed information is provided in the table below:

| | **Organism** | **Study pathway (PCR method), cases & controls** | **Clinical pathway (mixed methods), cases only** |
|---|---|---|---|
| Virus | 1 Influenza | Yes, also quantity | Yes |
| | 2 RSV | Yes, also quantity | Yes |
| | 3 HMPV | Yes, also quantity | Yes |
| | 4 Parainfluenza | Yes, also quantity | Yes |
| | 5 Adenovirus | Yes | Yes |
| | 6 Rhinovirus | Yes | Yes |
| | 7 Coronavirus (non-SARS-CoV-2) | Yes | No |
| Bacteria | 1 Streptococcus pneumoniae | Yes, also quantity | No |
| | 2 Haemophilus influenzae | Yes, also quantity | No |
| | 3 Moraxella catarrhalis | Yes, also quantity | No |
| | 4 Staphylococcus aureus | Yes, also quantity | No |
| | 5 Mycoplasma pneumoniae | Yes | Yes |

Are there any other organisms that you think important to consider for respiratory infection in children in Australia? If yes, please specify:

With regard to each organism listed above (V1-7, B1-5), along with the ones you have just specified, please answer the following questions:

- Where can the organism reside/be present **without** necessarily causing disease? Please delete sites as appropriate.

| **Organism** | **Sites in which presence is potentially harmless** | **Comment** |
|---|---|---|
| V1 Influenza | *Nasal passage/ ear/ throat/ lower airway/ None* | |
| V2 RSV | *Nasal passage/ ear/ throat/ lower airway/ None* | |
| V3 HMPV | *Nasal passage/ ear/ throat/ lower airway/ None* | |
| V4 Parainfluenza | *Nasal passage/ ear/ throat/ lower airway/ None* | |
| V5 Adenovirus | *Nasal passage/ ear/ throat/ lower airway/ None* | |
| V6 Rhinovirus | *Nasal passage/ ear/ throat/ lower airway/ None* | |
| V7 Coronavirus | *Nasal passage/ ear/ throat/ lower airway/ None* | |
| B1 Streptococcus pneumoniae | *Nasal passage/ ear/ throat/ lower airway/ None* | |
| B2 Haemophilus influenzae | *Nasal passage/ ear/ throat/ lower airway/ None* | |
| B3 Moraxella catarrhalis | *Nasal passage/ ear/ throat/ lower airway/ None* | |
| B4 Staphylococcus aureus | *Nasal passage/ ear/ throat/ lower airway/ None* | |
| B5 Mycoplasma pneumoniae | *Nasal passage/ ear/ throat/ lower airway/ None* | |
| *Please add as required* | *Nasal passage/ ear/ throat/ lower airway/ None* | |

| | |
|---|---|
| *Please add as required* | *Nasal passage/ ear/ throat/ lower airway/ None* |

- At each identified site above, how likely can each organism become pathogenic (infecting human cells at that site and causing inflammation): ***rarely, possibly, likely, very likely, always***? Please indicate how confident you feel about your overall responses for a given organism, e.g., low confidence=1 to high confidence=5, or leave a comment to elaborate on how easy or difficult it may be to provide a response. (Note that in all cases, your responses are very valuable to us.)

| **Organism** | **Nasal passage** | **Ear** | **Throat** | **Lower airway** | **Confidence/ comment** |
|---|---|---|---|---|---|
| V1 Influenza | | | | e.g. *always* | |
| V2 RSV | | | | | |
| V3 HMPV | | | | | |
| V4 Parainfluenza | | | | | |
| V5 Adenovirus | | | | | |
| V6 Rhinovirus | | | | | |
| V7 Coronavirus | | | | | |
| B1 Streptococcus pneumoniae | | | | | |
| B2 Haemophilus influenzae | | | | | |
| B3 Moraxella catarrhalis | | | | | |
| B4 Staphylococcus aureus | | | | | |
| B5 Mycoplasma pneumoniae | | | | | |
| *Please add as required* | | | | | |
| *Please add as required* | | | | | |

- If there is an infection at a given site (e.g., middle ear), what are the most likely pathogens to cause that infection?

| **Site of infection** | **Pathogen 1** | **Pathogen 2** | **Pathogen 3** | **Comment** |
|---|---|---|---|---|
| Nasal passage | | | | |
| Middle ear | | | | |
| Throat | | | | |
| Trachea/ bronchi | | | | |
| Alveoli/ bronchioles | | | | |
| Pleural space | | | | |

**Survey 3 – distributed prior to Workshop 3**

#### Survey - Overview

In the previous knowledge elicitation session in October 2020, we focused on identifying important pathogens that can cause infection in the respiratory tract. These pathogens include:

| Table 1. Key pathogens that can cause respiratory tract infection. | |
|---|---|
| **Viruses** | **Bacteria** |
| V1 Influenza | B1 Streptococcus pneumoniae |
| V2 RSV | B2 Haemophilus influenzae |
| V3 HMPV | B3 Moraxella catarrhalis |
| V4 Parainfluenza | B4 Staphylococcus aureus |
| V5 Adenovirus | B5 Mycoplasma pneumoniae |
| V6 Rhinovirus | B6 Strep pyogenes |
| V7 Coronavirus | B7 Pertussis |
| *Please add as required* | B8 Group B strep |
| | B9 Chlamydia pneumoniae |
| | *Please add as required* |

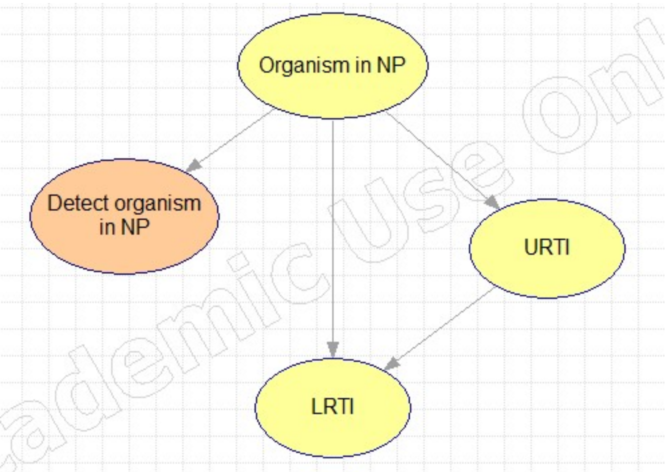

Figure 1. Model structure that illustrates the relationship between the presence of organism, infections and detection.

As illustrated in the figure above, given the presence of an organism in one's nasopharyngeal area (**organism in NP**), there is a chance for this organism to become pathogenic and infect the host's lower respiratory tract (causing **LRTI**). Infection of the LRT can either occur *directly* (the vertical arrow), potentially by direct aspiration/inhalation, or it can occur *indirectly* via secondary expansion from another site/s (e.g. secondary to **URTI**).

It is important to note that we explicitly distinguish between the actual presence of an organism at a body site (yellow nodes) from its successful detection by sampling that site (orange node), although we only ever know about the former via information about the latter. In the model, we treat the actual presence as a latent node, which means it cannot be observed directly, and this allows us to capture any factors that might affect the sensitivity and specificity of a detection method, such as age, specimen type, assay type, density of organism at the site and collection performance.

Figures 2-4 below show the PCR detection of different groups of organisms in children in PneumoWA with x-ray confirmed pneumonia (defined as *cases*; top row) and non-pneumonia *controls* (bottom row). The detection of bacteria in NP gradually decreases as age increases, but it does not differ between *cases* and *controls* (Figure 2). The detection of pathogenic viruses in the NP is strongly associated with being a case, while decreasing with age (Figure 3). The detection of less pathogenic viruses does not differ notably between cases and controls (Figure 4).

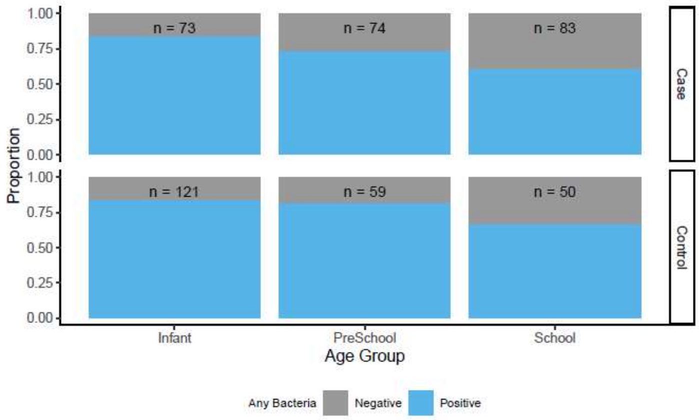

Figure 2. Bacteria refers to streptococcus pneumoniae, haemophilus influenzae, moraxella catarrhalis, staphylococcus aureus and/or mycoplasma pneumoniae (B1 – B5 in Table 1).

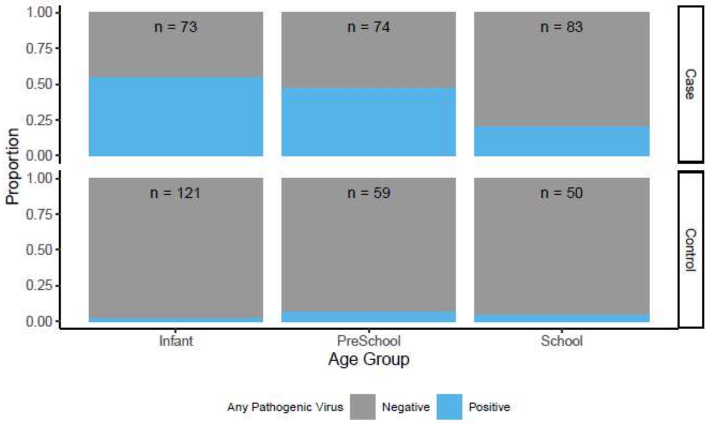

Figure 3. Pathogenic virus refers to influenza, respiratory syncytial virus, human metapneumovirus and/or parainfluenza (V1 – V4 in Table 1).

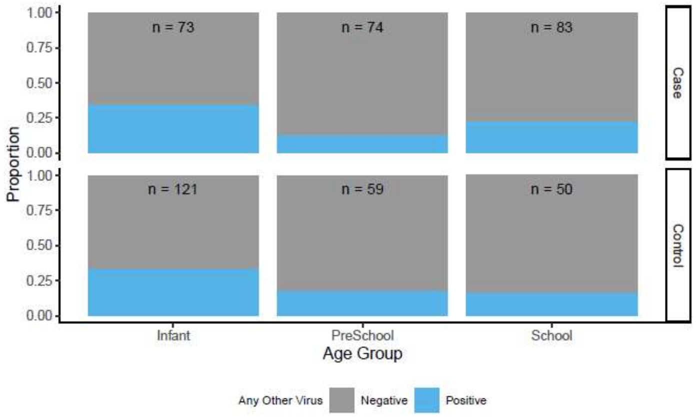

Figure 4. Other virus refers to adenovirus, rhinovirus and/or coronavirus (V5 – V7 in Table 1).

Figure 5 demonstrates how the varying levels of organism-specific pathogenicity can be described using models. The presence of bacteria, pathogenic virus and less pathogenic virus in NP are associated with 1.4%, 9.2% and 2.5% risk of pneumonia (these numbers are illustrative and *not* inferred by data).

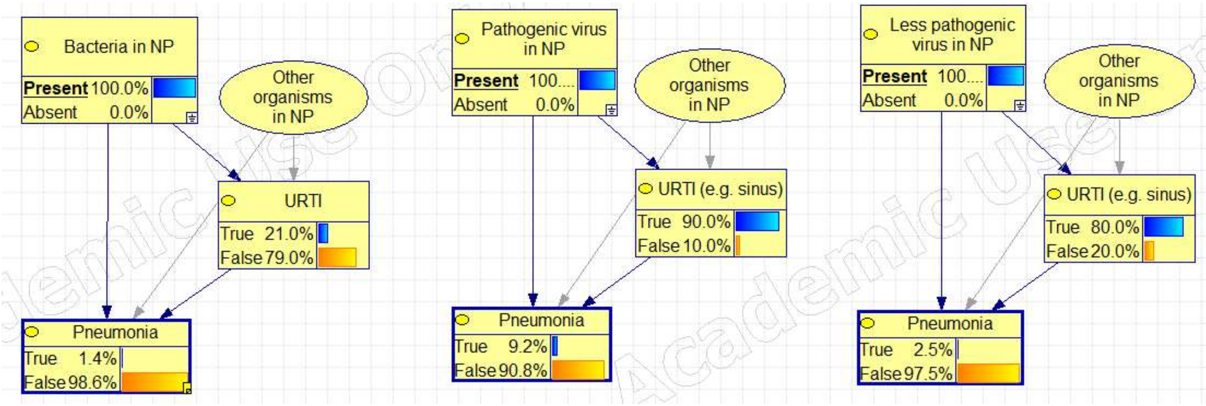

Figure 5. Preliminary models that illustrate varying levels of organism-specific pathogenicity

#### Questions

The following set of questions aims to better understand the pathogenicity of different organisms, and their subsequent implications on antibiotic use. Please provide your responses in the tables that follow, for each organism listed in the first column of the tables.

**1.** Imagine you randomly swab a child in the community without knowing anything about whether they have signs or symptoms of RTI. For each organism, order from highest to lowest your belief that its presence in the NP (whether detected or not) predicts that the child has (or will soon develop) a LRTI (which does not need to be caused solely by that specific organism). Please enter your order in the 2nd column, 1 for the highest and ties are allowed. Precise answers are not required. Provide your degree of confidence in your answer for each organism in the 3rd column, from highest 1 to lowest 5.

| **Bug X in NP** | **Q1. Order** *1 for the highest* | **Confidence** *highest 1 to the lowest 5* |
|---|---|---|
| V1 Influenza | | |
| V2 RSV | | |
| V3 HMPV | | |
| V4 Parainfluenza | | |
| V5 Adenovirus | | |
| V6 Rhinovirus | | |
| V7 Coronavirus | | |
| B1 Streptococcus pneumoniae | | |
| B2 Haemophilus influenzae | | |
| B3 Moraxella catarrhalis | | |
| B4 Staphylococcus aureus | | |
| B5 Mycoplasma pneumoniae | | |
| B6 Strep pyogenes | | |
| B7 Pertussis | | |
| B8 Group B strep | | |
| B9 Chlamydia pneumoniae | | |
| *Please add as required* | | |

**2.** Assuming a LRTI is diagnosed given the presence of the organism, what's the most likely type of causative pathogen/s for the LRTI (i.e., the pathogen/s causing inflammation of the LRT)? Please provide your answer in the 2nd column of the table below using *bacterial*, *viral*, or *both*.

**3.** For this LRTI, how likely is the patient going to benefit from antibiotic therapy? Please provide your answer in the 3rd column using: *high, moderate, or low.*

For example, the benefit of receiving antibiotic therapy might be *low* if the infection is caused by virus alone. The benefit might be *high* if the infection is bacterial and severe (e.g., with confirmed empyema). The benefit might be *moderate* if either the role of bacteria is unclear, or the infection is relatively mild and can possible self-resolve even if it's bacterial.

Precise answers are not required. Provide your degree of confidence in your answer for each organism in the last (4th) column, from highest 1 to lowest 5. (If you wish, you can also specify your confidence for specific questions.)

| **Bug X in NP** | **Q2. Most likely cause** *bacterial*, *viral*, or *both*. | **Q3. Benefit of antibiotics** *high, moderate, or low* | **Confidence** *highest 1 to the lowest 5* |
|---|---|---|---|
| V1 Influenza | | | |
| V2 RSV | | | |
| V3 HMPV | | | |
| V4 Parainfluenza | | | |
| V5 Adenovirus | | | |
| V6 Rhinovirus | | | |
| V7 Coronavirus | | | |
| B1 Streptococcus pneumoniae | | | |
| B2 Haemophilus influenzae | | | |
| B3 Moraxella catarrhalis | | | |
| B4 Staphylococcus aureus | | | |
| B5 Mycoplasma pneumoniae | | | |
| B6 Strep pyogenes | | | |
| B7 Pertussis | | | |
| B8 Group B strep | | | |
| B9 Chlamydia pneumoniae | | | |
| *Please add as required* | | | |

**4.** In Question 3, we provided some examples of reasons why antibiotics may or may not be beneficial. If you'd like to add further reasons or comment on the examples, please do so below:

#### Survey 4 – distributed prior to Workshop 4

The purpose of this survey is to evaluate the validity of the model (v1.3) based on expert opinion.

We present in the table below three hypothetical clinical scenarios (A, B, and C) where under each, clinical observations are entered (cumulatively) by steps (1, 2, etc.). According to information entered in each step, model-predicted outcomes get updated for all variables of interest, namely, the causative pathogen for pneumonia, positive detection of key pathogens in NP, the clinical diagnosis of pleural effusion, and pleural fluid and blood testing results.

The primary output of this model is a probability distribution over the causative pathogen for pneumonia, which can be bacterial, viral, or viral bacterial (mutually exclusive states). As a latent node, the causative pathogen cannot be observed and evaluated directly. Instead, a number of non-latent nodes can be used together to inform understanding of the model prediction. For example, as the predicted probability of viral pneumonia increases from its marginal 76% (in the cohort of 230 X-ray confirmed cases) to 84%, the predicted probability of RSV detection in NP also increases from the marginal 23% to 43% (demonstrated scenario A, step 1 in table below).

Please review the table and provide your comments on the following questions:

1. Do the model predictions look reasonable to you for each scenario and step? Please highlight the probabilities that do not meet your expectation and add a comment to explain why.
2. How useful do you think these model outputs would be if they were to be provided for clinical decision support?  What other information would you want to have for better diagnosis of the aetiology of pneumonia?
3. Please list three scenarios of your choice that you would put to the model to help you feel confident about the model's predictions. All variables included in the three figures in the appendix can be selected as either input or output variables.

**An example** of how to read the table. Among the 230 cases of X-ray confirmed pneumonia, on average, **4%** are *bacterial* infection, **20%** are *viral-bacterial co-infection*, and **76%** are *viral* infection based on the model. If more information were made available to the treating doctor, the model-predicted probability of bacterial, viral-bacterial and viral pneumonia would be updated:

A1) If the patient is a 4-month-old, with measured temperature of 37.8, runny nose, cough and diarrhoea: The predicted probability of *viral* infection increases from 76 to **83%**, while the probability of bacterial and viral-bacterial pneumonia drops to **2%** and **16%**, respectively.

A2) If for the same 4mo child, in addition to the above (A), it is also known that the child's CRP is above 70: The predicted probability for bacterial, viral-bacterial and viral pneumonia would update to **11%**, **27%** and **62%**, respectively.

A3) If in addition to all the above (A and B), it's further known to the clinician that the child's white cell count is above 18: The predicted probability for bacterial, viral-bacterial and viral pneumonia would again update to **24%**, **57%** and **19%**, respectively.

| Scenarios \ Model-predicted | | | **Causative pathogen for pneumonia** | | | **Positive detection in NP** | | | | | **Pleural effusion and blood** | | |
|---|---|---|---|---|---|---|---|---|---|---|---|---|---|
| | | | *Bacterial* | *Viral Bacterial* | *Viral* | **Mycoplasma** | **RSV** | **Flu** | **HMPV** | **Parainfluenza** | **Pleural effusion** | **+ve bac in tested PF** | **+ve bac in tested blood** |
| ***Baseline: marginal in 230 cases CXR confirmed pneumonia, all kids > 3mo present to PCH*** | | | **4.3%** | **20.2%** | **75.5%** | **14%** | **25.6%** | **11.7%** | **7.4%** | **6.2%** | **9.7%** | **44.1%** | **6%** |
| **A** | 1 | **4mo, temp 37.8, runny nose, cough and diarrhoea.** | **1.9%** | **15.6%** | **82.5%** | **7.9%** | **54.3%** | **10.3%** | **20.9%** | **5.1%** | **3.3%** | **52.3%** | **5%** |
| | 2 | The above plus CRP > 70 | 11% | 27.1% | 61.9% | 7.4% | 48.9% | 9.6% | 19.2% | 4.9% | 11.3% | 47.5% | 8.9% |
| | 3 | All the above plus WCC > 18 | 24.2% | 56.7% | 19% | 6.8% | 40.8% | 8.5% | 17.1% | 4.5% | 24.3% | 47.4% | 15.2% |
| **B** | 1 | **3.5yo, temp 39.5, vomiting and diarrhoea** | **10.3%** | **1.7%** | **87.9%** | **15%** | **28.8%** | **11.2%** | **17.4%** | **7.5%** | **12.5%** | **41.8%** | **5.7%** |
| | 2 | The above plus crackles, flu season (June), and 3 doses of SPN vaccine done. | 3.6% | 0.3% | 96% | 7% | 46.4% | 10.8% | 23.4% | 3.8% | 5.6% | 41.1% | 5.5% |
| | 3 | All the above plus wheezing | 2.3% | 1.2% | 96.5% | 7% | 50.3% | 10.4% | 25.2% | 3.6% | 3.5% | 41.4% | 5% |
| | 4 | All the above plus RSV NPA +ve | 0% | 0.4% | 99.6% | 3.5% | *Entered* | 7% | 10.8% | 3.9% | 0.9% | 39.3% | 4% |
| **C** | 1 | **9yo, resp rate >50, temp 37.9, runny nose, no breathing difficulty** | **14.2%** | **48.6%** | **37.2%** | **18.2%** | **22.3%** | **13.3%** | **18.5%** | **7.2%** | **17%** | **47.7%** | **10.2%** |
| | 2 | Same child, RR>50, but instead temp is 40, no runny nose, with breathing difficulty | 23.3% | 4.2% | 72.4% | 17.9% | 10.6% | 8.4% | 7.3% | 6.5% | 26.7% | 41.9% | 14.3% |
| | 3 | The above plus WCC > 18 | 51% | 5.4% | 43.6% | 9.5% | 6% | 5.5% | 3.7% | 4.5% | 51.2% | 42.2% | 26.4% |
| | 4 | WCC < 10 instead plus CRP < 30 | 0.8% | 3% | 96.2% | 26.1% | 14.9% | 10.7% | 10.9% | 7.9% | 7.1% | 39.5% | 4.8% |

**Survey 5 – parameterisation survey**

After workshop 4, due to the limitation of data (e.g., small sample size in certain subgroups) and the essential role of latent variables (e.g., those describe infection at the central part of the model), we decided to elicit all key parameters that are relevant to the latent variables to further improve the model.

A parameterisation survey that consists of questions on 35 variables was created, and divided into two sets (each covered 24 variables) with the aim to reduce the number of questions (burden) distributed to each expert. A 15-minute briefing session was delivered to clarify the purpose of the survey to each expert, in particular highlighting the following:

- Responses will be collated to form the priors of the BN, which will be later updated by the pneumoWA data. Therefore, precise answers are not required and qualitative estimates will be helpful to 'set up' the BN for latter parameterisation (learning from the data). E.g., 90% vs 10% rather than 68% vs 72%.
- You will receive a total of 24 questions (Set A or Set B):
    - Part I (13): clinical signs and symptoms
    - Part II (5): laboratory investigation
    - Part III (6): epidemiological context
- Example question types (see next page)

### Question type 1

By **viral-like pneumonia**, we mean a pneumonia that is *caused* by virus or Mycoplasma; and by **typical bacterial pneumonia**, we mean a pneumonia that is *caused* by typical bacteria.

#### Q1 Cough

**Defn** **Cough** can be directly driven by upper airway inflammation, irritation, or mucus production (e.g. postnasal drip from rhinorrhoea or croup) or can be from lower airways involvement with a pneumonia including the causative pathogen and its current severity.

**Scenario** Consider a child with **severe viral like pneumonia** and **no upper airway involvement**, what do you estimate the probability (min, max, best guess) that cough would be reported by parent during the emergency department presentation? Please note that the "min/max" should be plausible lower or upper values, not the extreme recordable value.

| Please estimate the probability of parental reported cough | **Min** | **Max** | **Best guess** |
|---|---|---|---|
| in % | | | |

**Scenario** Consider the above probability of cough as the baseline, how do the following factors may decrease or increase the baseline? Please estimate a scale relative to 1 for each question, e.g., x0.5, x1.2, x10, etc.

**Causative pathogen for pneumonia**

| | |
|---|---|
| *Viral-like* | 1 (baseline) |
| *Typical bacterial* | |

**Upper airway involvement**

| | |
|---|---|
| *None* | 1 (baseline) |
| *Nasopharynx only* | |
| *Throat only* | |
| *Both of the above* | |

Any further comments?

E.g., any other factors that may directly drive the presence of cough?

This is the end of Q1.

### Question type 1 (another example)

#### Q9 Age and pain

**Defn** The **presence of pain** (which might not be acertained) can be directly driven by inflammation of a specific body site, and the acertainment of pain is influenced by the age of child.

**Scenario** Consider a child with ***mild to moderate*** **chest pain** (presence), what do you estimate the probability (min, max, best guess) that chest pain would be noted for each age group? Please note that the "min/max" should be plausible lower or upper values, not the extreme recordable value.

| Please estimate the probability of noting an existing chest pain (in %) | **Min** | **Max** | **Best guess** |
|---|---|---|---|
| *>=5yo* | | | |
| *2 to 5yo* | | | |
| *<2yo* | | | |

**Scenario** Consider the above probabilities of acertaining pain as the baseline, how do the following factors may decrease or increase the baseline? Please estimate a scale relative to 1 for each question, e.g., x0.5, x1.2, x10, etc.

**Severity of inflammation**

| | |
|---|---|
| *Mild to moderate* | 1 (baseline) |
| *Severe* | |

**Potential relevant sites**

| | |
|---|---|
| *Chest pain* | 1 (baseline) |
| *Abdominal pain* | |
| *Ear ache* | |
| *Sore throat* | |
| *Headache* | |
| *Bodyache* | |
| *Back pain* | |
| *Joint pain* | |

### Question type 2

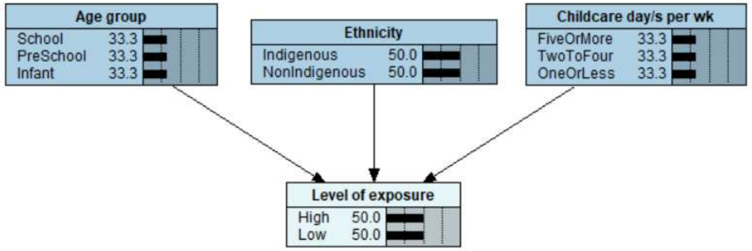

#### Q19 Level of exposure

**Defn**

The **level of exposure** refers to the child's exposure to pathogens with transmissible characteristics. In the model, it can be directly influenced by age, ethnicity, and the number of childcare days per week.

**Scenario**

Assume the following baseline level of exposure: a ***non-Indigenous***, ***less than 2-year-old*** child who attend **childcare for *<=1 day per week***. How do the following factors increase or decrease the exposure relative to the baseline? Please provide your min, max, and best guess estimates for each question. Note that the "min/max" should be plausible lower or upper values, e.g., 95th percentiles, not the extreme recordable value.

**Age**

| | | | |
|---|---|---|---|
| *<2yo* | Level of exposure = baseline | | |
| Please estimate a scale relative to 1, e.g., 0.5, 2 | **Min** | **Max** | **Best** |
| *2 to 5yo* | | | |
| *>=5yo* | | | |

**Ethnicity**

| | | | |
|---|---|---|---|
| *Non-Indigenous* | Level of exposure = baseline | | |
| Please estimate a scale relative to 1 | **Min** | **Max** | **Best** |
| *Indigenous* | | | |

### Question type 3

**Q14 CRP**

**Defn** Elevated level of C-reactive proteins (CRP) in blood can be driven by the systemic inflammatory response, which can be influenced by the causative pathogen for pneumonia and its current level of severtiy.

**Scenario** Consider a child with ***severe viral like* pneumonia**, what do you estimate the probability distribution for the level CRP?

| Please estimate the probability distribution of CRP | *<30* | *30 to 70* | *>=70* | |
|---|---|---|---|---|
| in % | | | | The row should sum to 100. |

**Scenario** Please estimate the above probability distribution for ***severe typical bacterial pneumonia*** .

| Please estimate the probability distribution of CRP | *<30* | *30 to 70* | *>=70* | |
|---|---|---|---|---|
| in % | | | | The row should sum to 100. |

Any further comments?

This is the end of Q14.

### Question type 4

22c About pathogenicity

**Scenario**

We have previously surveyed about the pathogenicity of different pathogens for causing lower respiratory tract infection LRTI (i.e., *causing LRTI given nasopharyngeal presence* ), and the average outcome order is provided in the table below (in column C, 1 for the highest). Please estimate the ratio of severe to mild infection if infection by a particular pathogen is confirmed.

| Pathogen | Pathogenicity | Ratio of severe to mild infection |
|---|---|---|
| *V1 Influenza* | 1.33 | |
| *V2 RSV* | 1.67 | |
| *V3 HMPV* | 3.17 | |
| *B7 Pertussis* | 3.67 | |
| *V4 Parainfluenza* | 3.83 | |
| *B5 Mycoplasma pneumoniae* | 4.33 | |
| *V5 Adenovirus* | 4.83 | |
| *V6 Rhinovirus* | 5.83 | |
| *B9 Chlamydia pneumoniae* | 6.50 | |
| *B1 Streptococcus pneumoniae* | 7.00 | |
| *B2 Haemophilus influenzae* | 7.00 | |
| *V7 Coronavirus (non-COVID)* | 7.17 | |
| *B3 Moraxella catarrhalis* | 7.50 | |
| *B4 Staphylococcus aureus* | 7.67 | |
| *B6 Strep pyogenes* | 7.67 | |
| *B8 Group B strep* | 8.83 | |

E.g., for every 1 child with severe lower airway infection with this pathogen, how many will have only moderate or mild infection (e.g., upper respiratory infection)?
